# The potential of SARS-CoV-2 antigen-detection tests in the screening of asymptomatic persons

**DOI:** 10.1101/2021.06.07.21258465

**Authors:** Jonas Wachinger, Ioana Diana Olaru, Susanne Horner, Paul Schnitzler, Klaus Heeg, Claudia M. Denkinger

**Affiliations:** Center of Infectious Diseases, Heidelberg University Hospital, Heidelberg, Germany; Clinical Research Department, Faculty of Infectious and Tropical Diseases, London School of Hygiene and Tropical Medicine, London, United Kingdom; German Center of Infection Research, Site Heidelberg, Heidelberg, Germany

**Keywords:** SARS-CoV-2, Ag-RDTs, screening, diagnostic yield, positive predictive value

## Abstract

**Objectives:** To assess the performance of antigen-based rapid diagnostic tests (Ag-RDTs) for SARS CoV-2 when implemented for large-scale universal screening of asymptomatic individuals.

**Methods:** This study presents data from a pragmatic implementation study for universal Ag-RDT-based screening at a tertiary care hospital in Germany where all incoming patients without symptoms suggestive of SARS-CoV-2 were screened with an Ag-RDT prior to admission since October 2020.

**Result:** In total, 49,542 RDTs were performed in 27,199 asymptomatic individuals over a duration of five months. Out of 222 positive results, 196 underwent in-house confirmatory testing with PCR, out of which 170 were confirmed positive, indicating a positive predictive value (PPV) of 86.7%. Negative Ag-RDTs were not routinely tested with PCR, but a total of 94 cases of false negative Ag-RDTs were detected due to PCR tests being performed within the following five days with a median CT-value of 33.

**Conclusions:** This study provides evidence that Ag-RDTs can have a high diagnostic yield for transmission relevant infections with limited false-positives when utilized at the point of care on asymptomatic patients and thus can be a suitable public-health test for universal screening.

## Introduction

Several researchers and policy makers, supported by evidence from modelling studies, have recently argued to increase large-scale screening for SARS-CoV-2 to curb transmission from patients with minimal or no symptoms.[1–5] Antigen-detection point-of-care rapid diagnostic tests (Ag-RDTs) have shown very good sensitivity (88%) in persons with high viral load (CT <30) along with high specificity (>99%).[6, 7] With their favourable ease-of-use, rapid turn-around, and good (although suboptimal) performance, Ag-RDTs meet the characteristics for a test for public health use and could allow for better control of transmission if implemented in well-designed universal screening strategies.[8, 9] However, one frequently raised concern has been the potentially insufficient specificity, leading to large numbers of false-positives when using Ag-RDTs in large-scale screening strategies with low prevalence, which could conceivably disrupt workflows and undermine trust in the test. Furthermore, in a setting where high-risk persons are present (e.g., hospital), concerns exist regarding imperfect sensitivity leading to secondary cases and substantial morbidity and mortality. Data from large scale screening implementation efforts that would allow to gauge diagnostic yield and issues with false-positives are limited.

## Methods

We performed a large-scale, pragmatic implementation study of Ag-RDTs in the context of a universal screening program at one of Germany’s largest tertiary care hospitals (Heidelberg University Hospital, Germany) serving over 100,000 inpatients and 1.3 million outpatients per year.[10] The study was conducted between 20 September 2020 and 7 March 2021. Patients without SARS-CoV-2 associated symptoms presenting for elective procedures or outpatient treatment requiring close contact or longer presence (e.g., dialysis) were screened with an Ag-RDT. Depending on setting and local SARS-CoV-2 infection dynamics, other external personnel (e.g., craftspeople, visitors, translators) were similarly screened. Trained nursing staff performed the STANDARD Q COVID-19 Ag Test (SD Biosensor, Inc. Gyeonggi-do, Korea), a WHO recommended and independently validated instrument-free lateral flow assay for SARS-CoV-2 detection,[11] using nasopharyngeal swabs. In patients with a positive Ag-RDT, an additional nasopharyngeal swab was collected for confirmatory SARS-CoV-2 PCR. Ag-RDT results were confirmed with PCR in selected departments prior to ward admission (e.g., haematology), prior to planned procedures associated with high levels of aerosol production, or when a patient developed SARS-CoV-2 associated symptoms or a cluster of cases occurred. To analyse diagnostic yield of Ag-RDTs and false positives, we systematically extracted results of the Ag-RDTs, as well PCR-tests (with CT values) that were done within 5 days after Ag-RDT screening. PPV and sensitivity were computed using the confirmatory PCR result as reference standard. Analysis was conducted using R v4.0.3 (The R Foundation). The ethical review board at Heidelberg University approved this study (S-811/2020). PPV and sensitivity were computed using the confirmatory PCR result as reference standard.

## Results

Between 20 September 2020 and 7 March 2021, 49,542 Ag-RDTs were performed in 27,421 asymptomatic individuals. Ag-RDTs were positive in 222 individuals and 49,320 Ag-RDTs were negative in 27,199 individuals. Out of the 222 individuals with positive Ag-RDTs, 196 (88.3%) were also tested using PCR in-house. The PPV for the Ag-RDTs was 170/196 (86.7%, 95%CI 81.2-91.1%). Among patients with a positive confirmatory PCR performed within 5 days of a positive Ag-RDT, the median CT value was 19 (IQR 15-24, Figure 1). Of 27,421 patients with a negative Ag-RDT, 94 had a positive PCR in the 5 days following the Ag-RDT. The median number of days between the tests was 1 (IQR 0-1). Based on these false-negative cases identified via PCR, the overall sensitivity of the Ag-RDT can be estimated to be 170/264 (64.4%, 95%CI 58.3-70.2). The median CT value of patients who were missed using Ag-RDTs was 33 (IQR 29-35). In total, only 12/94 (12.8%) Ag-RDT false-negative patients had a PCR with a CT-value <25, and 10/12 were identified on the same or following day.

**Figure 1.**
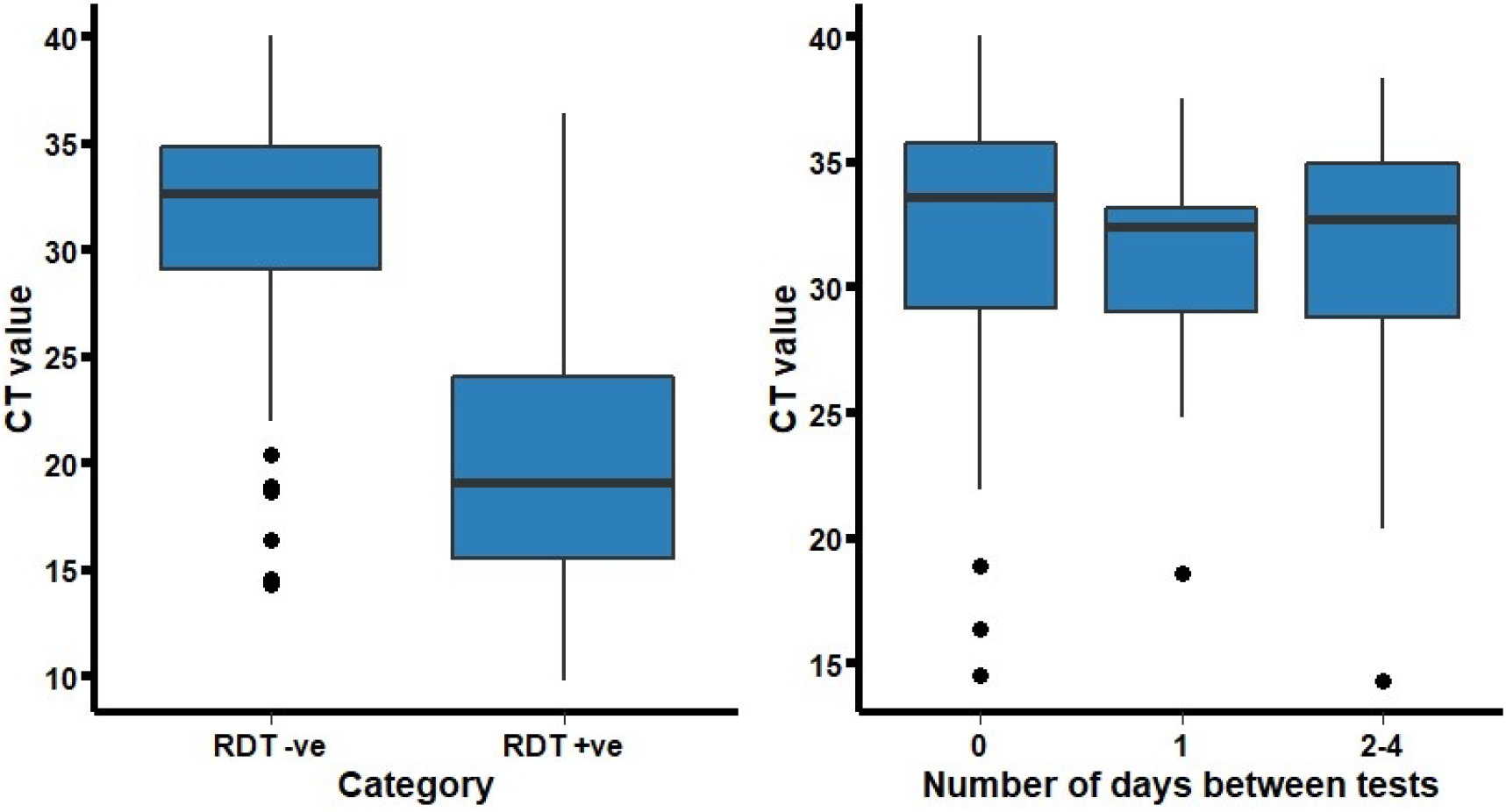
SARS-CoV-2 PCR CT-values according to Ag-RDT result (left) and for false-negative Ag-RDT results according to the number of days between discordant tests (right).

## Discussion

This pragmatic implementation study of a universal screening programme of asymptomatic patients at a tertiary care hospital showed the benefit of Ag-RDTs identifying SARS-CoV-2 infected persons who would have otherwise entered a high-risk environment leading to potential secondary transmission. The sensitivity observed in this study is higher than that observed in other studies of asymptomatic infections.[7] Although the data on accuracy in asymptomatic patients is limited, we acknowledge that the sensitivity estimate is likely an overestimate, as not all patients with asymptomatic infections were PCR-tested in this pragmatic study. However, the study confirms that most cases at high-risk of causing secondary infections are detected via Ag-RDT. Missed cases with CT<25 on PCR testing were mostly captured within 24hrs and are likely attributable to a negative Ag-RDT in the early phase of disease when the viral load is increasing rapidly.[12] Furthermore, the study showed a very high PPV of the Ag-RDT, thus confirming the high reliability of a positive result on an Ag-RDT shown in accuracy studies.[7]

To the best of our knowledge, this is the first large-scale implementation study of a universal screening program of asymptomatic individuals to analyse the diagnostic yield of Ag-RDTs. The central limitation of this study, inherent to a pragmatic implementation study, is the limited confirmatory PCR testing for negative Ag-RDTs.

In conclusion, this study provides evidence that an Ag-RDT can be a suitable test for large-scale universal screening in a hospital setting and add the important component of a public-health test to our diagnostic armamentarium.

## Data Availability

Datasets used and/or analysed during the current study are available from the corresponding author on reasonable request. However, to preserve the anonymity of respondents and considering the personal nature of this data, requests will be considered on a case-by-case basis.

## Acknowledgments

None

## Author’s Contributions

JW, KH, and CMD conceptualized the study. IDO analysed the data, supported by JW, SH, PS, and CMD. JW, IDO, and CMD drafted the manuscript. All co-authors critically revised and approved the final version of the letter.

